# Identifying Patterns of ALS Progression from Sparse Longitudinal Data

**DOI:** 10.1101/2021.05.13.21254848

**Authors:** Divya Ramamoorthy, Kristen Severson, Soumya Ghosh, Karen Sachs, Answer ALS, Jonathan D. Glass, Christina N. Fournier, Pooled Resource Open-Access ALS Clinical Trials Consortium, James Berry, Kenney Ng, Ernest Fraenkel

## Abstract

Amyotrophic Lateral Sclerosis (ALS) is a neurodegenerative disease that is complex in its onset, pattern of spread, and disease progression. The heterogeneity of ALS makes it extremely challenging to determine if a disease modifying therapy is effectively slowing progression. While accurately modeling ALS progression is critical to developing therapeutics, current computational methods fail to capture the complexity of disease progression. We aimed to robustly characterize disease progression patterns in ALS.

We obtained data from four clinical cohorts that cover more than 3,500 patients and include both observational and clinical trial studies. To determine whether there were common patterns of disease progression, we developed an approach based on a Mixture of Gaussian Processes (MoGP) to model longitudinal clinical data. Our approach automatically identifies clusters of patients who show similar disease progression patterns, modeling their average trajectory and the spread of the distribution in each cluster. Importantly, the method does not require any prior knowledge of the expected number of clusters.

The MoGP approach revealed that ALS progression, as measured using the ALS functional rating scale (ALSFRS-R) or forced vital capacity, is often non-linear with periods of stable disease preceded or followed by rapid decline. Patterns of progression in ALSFRS-R were robust to sparse data. When at least one year of longitudinal data were available, MoGP predictions were significantly more accurate than linear models, which are commonly used in clinical trials. Progression patterns were consistent across different cohorts despite differences in the frequency of data collection and the lengths of follow-up periods. We further showed that clusters identified from one large, publicly available study population could be used to stratify unseen participants in other studies. We also showed that these progression trajectories correspond with survival outcomes.

This work highlights the importance of modeling nonlinear disease progression for developing more advanced clinical trial endpoint analysis models. In ALS, sporadic, rapid decline (“functional cliffs”) and sigmoidal patterns in disease progression in untreated patients may obscure detection of therapeutic efficacy if linear models are used. We provide a pre-trained computational model of observed clinical patterns that can be used by others to analyze new ALS patient cohorts. We expect that the MoGP approach can also be applied to additional ALS outcome measures and to other progressive diseases. Our results provide a critical advance in characterizing the complex disease progression patterns of ALS.

## Introduction

Amyotrophic Lateral Sclerosis (ALS) is a neurodegenerative disease with a complex pathophysiology resulting in heterogeneous symptoms and progression.^1,2^ The median length of survival from symptom onset is approximately three years; however, some patients survive decades with the disease.^3^ The heterogeneity of ALS progression makes it extremely challenging to determine if a disease modifying therapy is effectively slowing progression.^4,5^

Despite widespread use of longitudinal functional clinical metrics to determine ALS progression, they are imperfect measures. Patients often are evaluated at different stages of their disease.^6^ While patients with ALS invariably decline over time, some of the metrics can increase for short durations or reach plateaus.^7^ Additionally, the metrics of clinical disease progression are based on subjective assessments of functional ability, such as the ability climb stairs “normally” or “slowly”, which introduces a potential source of error.^8^ The interconnectedness of function and variability in the measurement of these clinical metrics present challenges in modeling ALS progression.

Accurately modeling progression of ALS is critical to developing therapeutics. Traditional modeling approaches have dealt with the complexities in ALS clinical scores by first assuming that ALS outcome measures, particularly the Revised ALS Functional Rating Scale (ALSFRS-R), progress in a linear fashion.^9–11^ Many ALS clinical trials use changes in the linear slope of ALSFRS-R over time or a change in ALSFRS-R from baseline as primary endpoints.^12–14^ Relatively small improvements in the linear rate of decline of the ALSFRS-R are assumed to correspond with clinically meaningful efficacy. For example, edaravone was approved based on a 2.5 ALSFRS-R point difference in decline between the treatment and control arms over 6 months^14^, and the estimated effect from the ALS Sodium Phenylbutyrate–Taurursodiol clinical trial was a change in slope of 0.42 points per month.^12^ Large global crowdsourcing analyses designed to produce better models for clinical trials have also assumed a linear decline in ALSFRS-R.^15,16^

Despite the widespread use of linear models in predicting patient progression, there is evidence that ALS progression can be nonlinear and can differ across disease severity.^17–19^ Nonlinear parametric models that assume a particular shape of the trajectory in advance have been used to capture these complexities. However, by requiring a particular parametric form, these models are restricted to identifying pre-specified trajectory shapes^17,18,20–22^ or pre-specified subgroups^19,23^, which may not represent the actual heterogeneity in disease progression patterns.

To model the full complexity of ALS progression, we turned to computational methods that are more flexible than traditional parametric methods. We propose a framework for aggregating patient trajectories into trajectory clusters by determining the overall shape of the trajectories in each cluster using Gaussian processes^24,25^, and determining the number of clusters using a Dirichlet process mixture model.^26,27^ Gaussian process models make minimal assumptions of expected trajectory shapes, while flexibly allowing for the incorporation of prior clinical knowledge. For example, since patients with ALS are expected to decline over time, we incorporate monotonic biases into our model, which encourage declining trajectories to be identified but also allow for the detection of patterns that do not fit prior expectations. Dirichlet process mixture models do not require the specification of the number of clusters in advance and instead learn the number of clusters from the data. Our approach accounts for noise in clinical metrics, both providing an estimate of predicted cluster membership for each patient and also providing an estimate of error around each cluster.

We show that this method can improve characterization of ALS progression patterns, and identify clusters of participants with similar trajectories from longitudinal clinical scores. We show that nonlinear progression patterns are robust to sparse data, consistent across study populations, and correspond with survival outcomes. While we focus on clinical ALS outcome measures, this framework can be applied to any longitudinal measure used in ALS or in other diseases. Our results provide critical advances in modeling ALS progression patterns.

## Methods

### Study Populations

Data on longitudinal ALSFRS-R scores was obtained from four study populations (Table 1). Two observational studies were used: Answer ALS (AALS)^28^, and the Emory ALS Clinic database (EMORY)^3^, and two overlapping clinical trial datasets: The Pooled Resource Open-Access ALS Clinical Trials (PRO-ACT)^29^ and the Clinical Trial of Ceftriaxone in ALS (CEFT).^13^ The heterogeneity of the clinical cohorts enabled us to measure the robustness of our model between the study populations.

**Table 1:** Study Populations. Abbreviations: PRO-ACT = Pooled Resource Open-Access ALS Clinical Trials; AALS = Answer ALS; CEFT = Clinical Trial Ceftriaxone in Subjects With ALS; EMORY = Emory ALS Clinic

| Dataset | Study Type | Total No.<br>Participants<br>Included | Number Visits<br>Mean (SD) | Months<br>Followed<br>Mean (SD) | ALSFRS-R<br>Slope<br>Mean (SD) |
| --- | --- | --- | --- | --- | --- |
| PRO-ACT | Clinical Trial | 2923 | 10 (4) | 11.86 (6.33) | -1.09 (0.99) |
| CEFT | Clinical Trial | 476 | 10 (5) | 19.09 (10.58) | -1.20 (0.79) |
| AALS | Observational | 456 | 5 (2) | 16.64 (8.78) | -0.75 (0.65) |
| EMORY | Observational | 399 | 6 (4) | 19.47 (14.74) | -1.21 (1.20) |

### Modeling Approach

We developed a Mixture of Gaussian Processes model with strong inductive bias towards monotonic decline (MoGP) to characterize patterns in disease progression. The model leverages two Bayesian nonparametric methods: Gaussian process regression^24,25^ and Dirichlet process clustering.^26,27^ Gaussian process regression does not require the specification of a particular functional form, but instead learns trajectories from data, enabling the model to capture a wide variety of, possibly nonlinear, progression patterns. Dirichlet process clustering does not require the specification of a number of clusters a priori, but instead proposes a number of clusters that is consistent with the number of trajectory trends observed in the data. The number of patients in each cluster is also learned from the model, and clusters can differ in size from each other. This is well motivated by uncertainty in the existence and number of ALS progression subtypes and avoids restrictive modeling assumptions. In contrast to prior work on MoGP models,^25,30,31^ significant modifications to the method have been made to ensure that clinical knowledge relevant to ALS progression is incorporated. This includes the implementation of a monotonic inductive bias, as well as clinically-informed parameter priors for Gaussian process regression and Dirichlet process clustering components. Each component of the model is discussed in more detail below.

### Gaussian Process Regression

Gaussian process regression allows the identification of nonlinear trajectory patterns while making minimal assumptions about the shape of the trajectory functions.^24,25^ A Gaussian process is specified by a mean function and a covariance kernel. Because we expect ALS trajectories to be smooth functions with no discontinuities, our MoGP model uses a squared exponential (SE) kernel. The SE kernel has two parameters: the signal variance, which determines the average distance of the function from the mean and the length-scale, which specifies the smoothness of the function. Each of these parameters is determined during the learning phase using the training data.

### Monotonic Inductive Bias

Because ALS trajectories are expected to decline over time, we use a negative linear function in the Gaussian process models of MoGP, as well as a threshold function with our clustering to encourage declining trajectories. We also impute an onset-anchor value, a maximum score of a clinical metric assigned to the date corresponding to symptom onset, which has been previously shown to improve prediction in ALS trajectories.^32^

### Dirichlet Process Clustering

Dirichlet process mixtures^26,27^ can be used to identify clusters in data in cases in which it would be difficult to specify an expected number of clusters *a priori*, as in ALS subtyping. This unsupervised learning model begins by assuming that an infinite number of clusters can exist, and then narrows its prediction to a limited number of components best supported by the observed data. In our case, each mixture component is a function drawn from a Gaussian process. The resulting Dirichlet process mixture of Gaussian processes clusters patient trajectories by probabilistically assigning them to those components that best explain them. The number of patients in each cluster is also learned from the model, and clusters can differ in size from each other. Through this data-driven approach, the algorithm can learn clusters of ALS patients that share disease progression patterns. The method can also predict the cluster membership and the disease progression pattern of a participant not included in the model, and provide an estimate of the confidence of that prediction. For further details, please see the methods section of the supplement.

## Model Evaluation

### Evaluating trajectory nonlinearity

We compared the performance of MoGP against a calculated slope of each individual participant’s ALSFRS-R score (Slope Model: SM). The SM represents a common standard model in the field used to represent ALS disease progression, in which linear slopes are fit to patient data with an onset-anchor.^32^ The slope model is fit to each patient and does not identify clusters. We also benchmarked our model against a Mixture of Gaussian Processes model with a linear kernel (Linear Kernel Model: LKM). This baseline retains the ability to cluster trajectories using a Dirichlet process but does not allow for nonlinear functions, allowing us to separate the contribution of clustering and the assumption of linearity in our models.

For this analysis, participants were excluded from the model if fewer than three complete ALSFSR-R visits were recorded, the first visit was more than seven years from symptom onset, or an increase of greater than six points in ALSFRS-R between subsequent visits was recorded (Table 1).

For each model, the root mean squared error (RMSE) between a participant’s measured scores and their predicted cluster model mean function were calculated. The RMSE was compared between the models; a lower RMSE indicates reduced error in that model and better model performance.

### Robustness to Sparse Data

To evaluate the robustness of the MoGP cluster assignments, we simulated sparsity by withholding data and assessed the model’s ability to perform two tasks: 1) interpolation of ALSFRS-R scores (“Interpolation”) for a patient with randomly withheld data points, and 2) forecasting future ALSFRS-R disease progression (“Prediction”) for patients with right-censored data.

We tested this using the largest dataset with sufficient longitudinal measurements: PRO-ACT. We also evaluated the model’s performance on CEFT, which is a clinical trial that is a subset of the PRO-ACT study population. Including CEFT allows us to characterize the contribution that data collection heterogeneity and length of follow-up have on the results. In order to have sufficient longitudinal measurements, for Interpolation experiments, we only included participants with 10 or more longitudinal ALSFRS-R visits, and for Prediction experiments, we only included participants with 4 or more visits (Supplementary Table 1).

The reconstruction error for each participant was calculated using the RMSE between the original, withheld data points and predicted values from the mean function for the participant’s trajectory cluster. This was done across all interpolated tests, in which 25%, 50%, and 75% of clinic visits per patient were provided as training data, with selections randomly interspersed across visits. We then evaluated the ability of MoGP to predict future progression, by using right-censored data with varying amounts of training data (including visits within 0.25, 0.5, 1.0, 1.5, and 2.0 years since baseline visit).

### Model Generalizability

To evaluate whether clusters derived from one study population could be used to model external study populations, we trained a reference model and evaluated the transferability of this model to unseen ALS patient data. We predicted the cluster membership for each participant, and calculated the RMSE between the participant ALSFRS-R scores and the mean function of their predicted cluster.

We split all of our study populations into test and training datasets (60% train, 40% test). For our reference MoGP model, we used the training data from PRO-ACT, which was chosen because it contained the largest number of samples and is publicly available. For AALS, EMORY, and CEFT, we used the training data from each study to train a separate model (“study-specific model”). For each study’s remaining test data, we predicted the trajectory function using the reference model and the study-specific model.

### Relationship to Alternate Outcome Measures

We calculated the Kaplan-Meier survival probability curves for the largest MoGP clusters identified from PRO-ACT. If no death was recorded, the participant was marked as censored using the latest date of recorded ALSFRS-R score. We also trained MoGP models on forced vital capacity maximum percentages and ALSFRS-R subscores, and evaluated trajectory patterns.

### Statistical Analysis

For comparing the cumulative distribution function of the RMSE between a participant’s predicted cluster membership and cluster model mean, P-values calculated with Kolmogorov-Smirnov two-sample tests. For Interpolation and Prediction, to compare if a model error had decreased between the LKM or SM to the MoGP, a Wilcoxon signed-rank one-sided test was used. For assessing trajectory consistency between reference models and study-specific models, a Wilcoxon signed-rank two-sided test was used.

## Data availability

We provide the python code for the MoGP framework as well as the pre-trained reference model described here for researchers to use to generate predictions of cluster membership and trajectory function from input patient data. All code used for data processing, modeling, and figure generation can be found at: https://github.com/fraenkel-lab/mogp

AALS is publicly available for download (data.answerals.org). PRO-ACT can be downloaded by request (https://nctu.partners.org/ProACT). CEFT can be downloaded from National Institute of Neurological Disorders and Stroke (NINDS) (https://www.ninds.nih.gov/Current-Research/Research-Funded-NINDS/Clinical-Research/Archived-Clinical-Research-Datasets) by request. EMORY is restricted access at this time.

## Results

### Study populations

The populations varied in size, with PRO-ACT, a publicly available ALS clinical trial database, having the largest total number of participants (2923 participants with at least 3 ALSFRS-R visits recorded). The populations differed significantly in the average number months followed (between 11 to 20 months), and the average frequency of clinical visits (between 5 to 10 visits). The slope between the populations also varied, with EMORY on average having the fastest progressing population (−1.21 ALSFRS-R points/month), and AALS having the slowest progressing population (−0.75 ALSFRS-R points/month). CEFT had an average of 19.09 months of follow-up, while PRO-ACT had an average of 11.86 months, indicating that CEFT participants likely comprise some of the longest subject-records in PRO-ACT. The heterogeneity of the populations enabled us to measure the robustness of our model to data collection methods and the generalizability of ALS progression patterns between varying study populations.

### ALS disease progression trajectory patterns can be characterized with MoGP

MoGP is a flexible modeling and clustering framework that can be applied to linear and non-linear data. To characterize patterns in ALS progression, we first applied MoGP to PRO-ACT, which is the largest publicly available dataset of ALSFRS-R scores (Fig. 1). The analysis identified diverse clusters, including some clusters identifying slow progressing populations (Fig. 1N mean ALSFRS-R slope = -0.13 points/month) and others capturing faster progressing groups (Fig. 1R, mean ALSFRS-R slope = -1.93 points/month).

**Figure 1.**
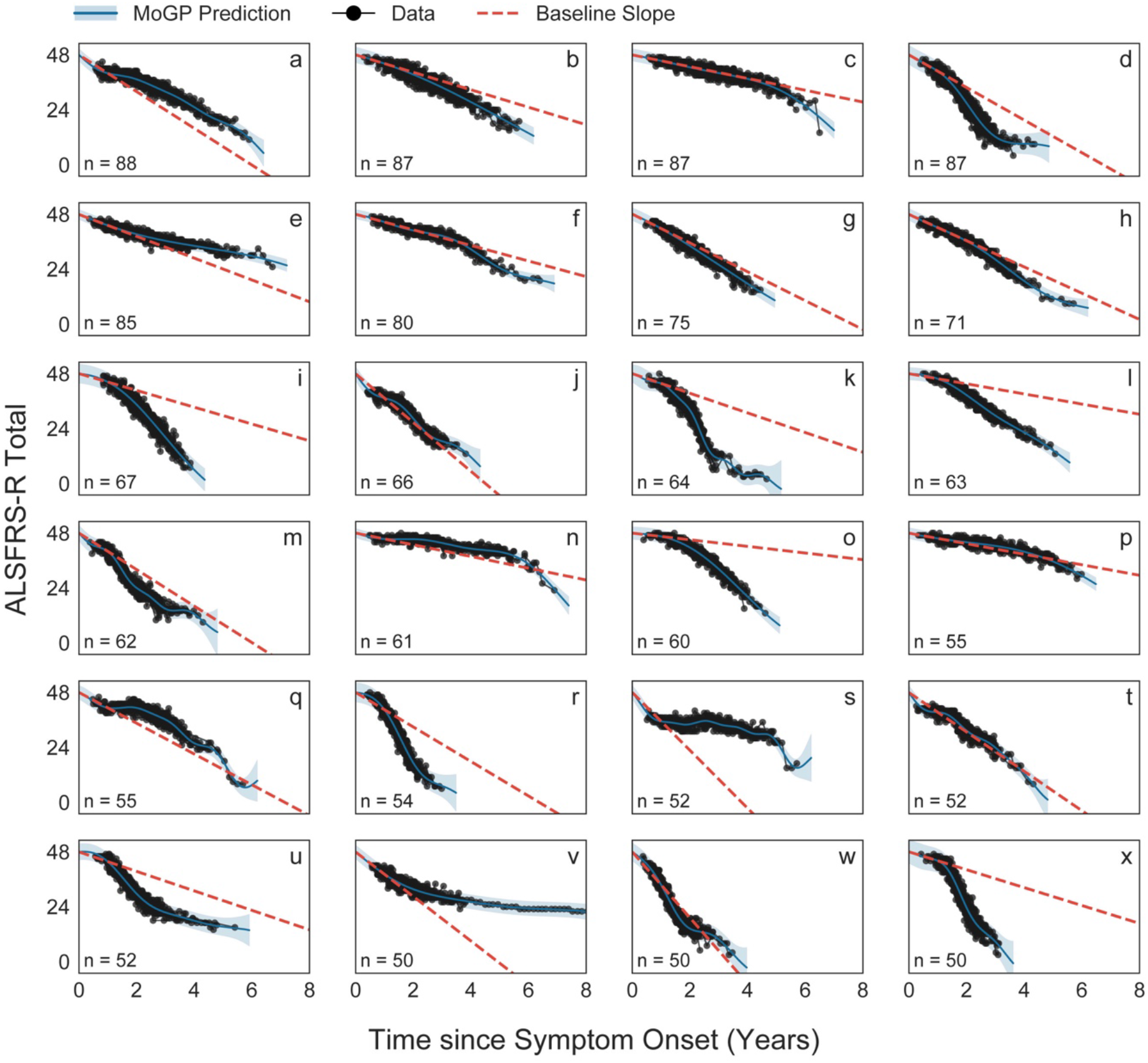
A Mixture of Gaussian Processes model identifies trajectory clusters with varying patterns of decline. The 24 largest clusters (out of 92) from PRO ACT are visualized. The baseline slope is calculated as the difference between 48 and the mean cluster score one year after symptom onset. N indicates the number of ALS patients in each cluster.

Notably, in many cases, the patterns of decline were highly non-linear, with some following sigmoidal (Fig. 1D, K), convex (Fig. 1M, U, V), and concave (Fig. 1O, Q) curves. Linear patterns were also detected in some clusters (Fig. 1G, J, T). To estimate the extent of non-linearity, we computed the slope in the first year since symptom onset (“Baseline slope”) for each cluster trajectory. While this baseline slope closely reflects actual trajectories for the linear clusters (Fig. 1G, J, T), for others, the baseline slope is either an overestimation (Fig. 1I, K, L, O, X), or underestimation (Fig. 1S, V), indicating nonlinearity in the trajectory pattern. These errors in baseline estimations can be large; for instance, the baseline slope in cluster K overestimates disease trajectory by 24.20 ALSFRS-R points and underestimates trajectory in cluster V by 9.48 ALSFRS-R points when both are evaluated 3 years after symptom onset. This diversity highlights the complexity of progression trajectories in ALS. Analysis of other study populations (Supplementary Fig. 1) also revealed many clusters that were highly non-linear.

### Nonlinear disease progression patterns are identified across heterogeneous study populations

To rigorously compare linear and non-linear models, we compared our MoGP against two benchmark linear models, described in the methods section: a slope model (SM) and a Mixture of Gaussian process model with a linear kernel (LKM).

For all study populations, the error in the MoGP model was lower than the LKM and SM (Fig. 2A). Across the populations, using non-linear models results in a lowered error of one or more ALSFRS-R points as compared to the LKM for at least 27.16% of participants; at least 8.33% of patients have an improvement in accuracy of two or more ALSFRS-R points (Supplementary Table 2). Importantly, the error of the MoGP was lower even though the LKM used a larger number of clusters to model the data (Supplementary Table 4). It is also notable that the MoGP, which identified clusters as large as 100 participants, was able to match or outperform the patient-specific SM (Fig. 2A, Supplementary Table 3), which would have been expected to drastically outperform MoGP if significant nonlinear structure did not exist in the data. The results are replicated across the four different datasets, suggesting that complex nonlinearity is a common feature of ALS progression, and is not a unique feature of a single dataset.

**Figure 2.**
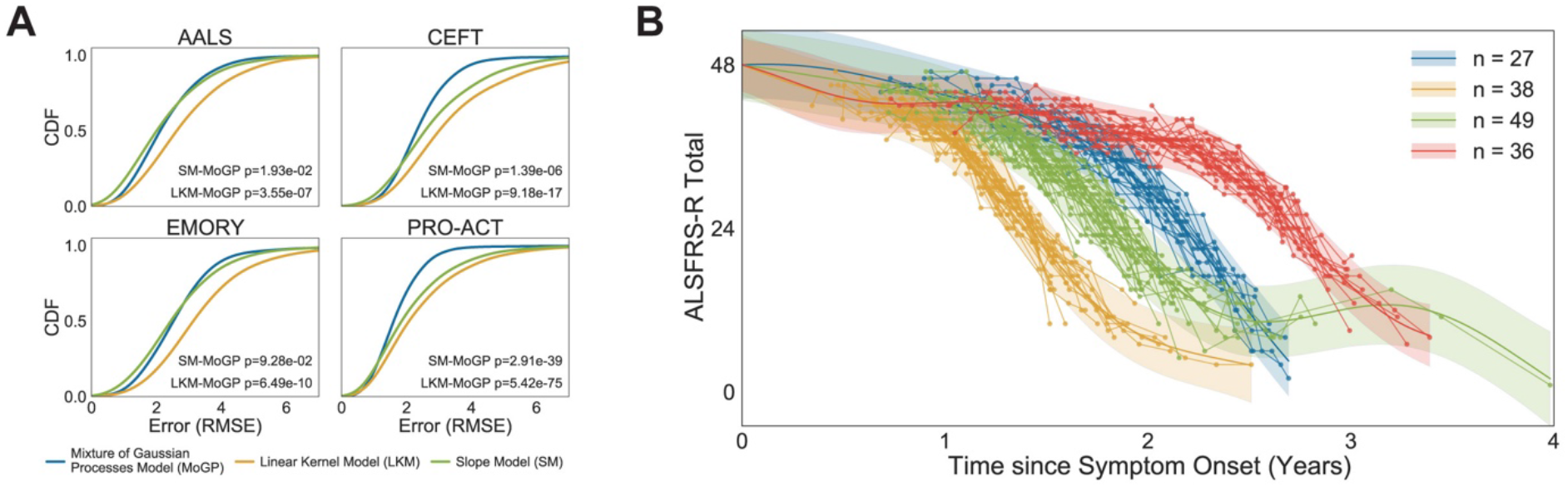
Estimating nonlinearity of trajectories. (A) Cumulative distribution function (CDF) of RMSE between a participant’s predicted cluster membership and cluster model mean. P-values calculated with Kolmogorov-Smimov two-sample tests between MoGP and LKM distributions, and between MoGP and SM distributions (B) A subset of nonlinear clusters from PRO-ACT Visualized: N indicates number of ALS patients per cluster

The clusters with the most significant nonlinearity often followed sigmoidal trajectory patterns, with varying inflection points (Fig. 2B). In some of these clusters, patients had slow progression for a period of time, followed by a consistent sharp decline. This pattern of progression appears consistent with a sudden loss of ability to carry out functions that we refer to as a “functional cliff.” In other cases, the pattern is more consistent with a rapid period of decline followed by a slower phase. The MoGP model enables the ability to learn these complex disease progression trajectories.

### Disease progression trajectory clusters are robust to sparse data

Clinical data for ALS patients can often be incomplete or sparse, and we sought to evaluate MoGP performance in these settings. We compared MoGP’s performance against the Linear Kernel Model (LKM) and the Slope Model (SM). Across all interpolated tests for PRO-ACT, we found that the clusters identified by MoGP had lower reconstruction error than the LKM (Fig. 3A, *p-val* ≤ 1e-4), and a lower error than the SM when 50% and 75% of training data is included (Fig. 3A, *p-val* ≤ 1e-3). These trends persisted when compared to CEFT (Fig. 3C).

**Figure 3.**
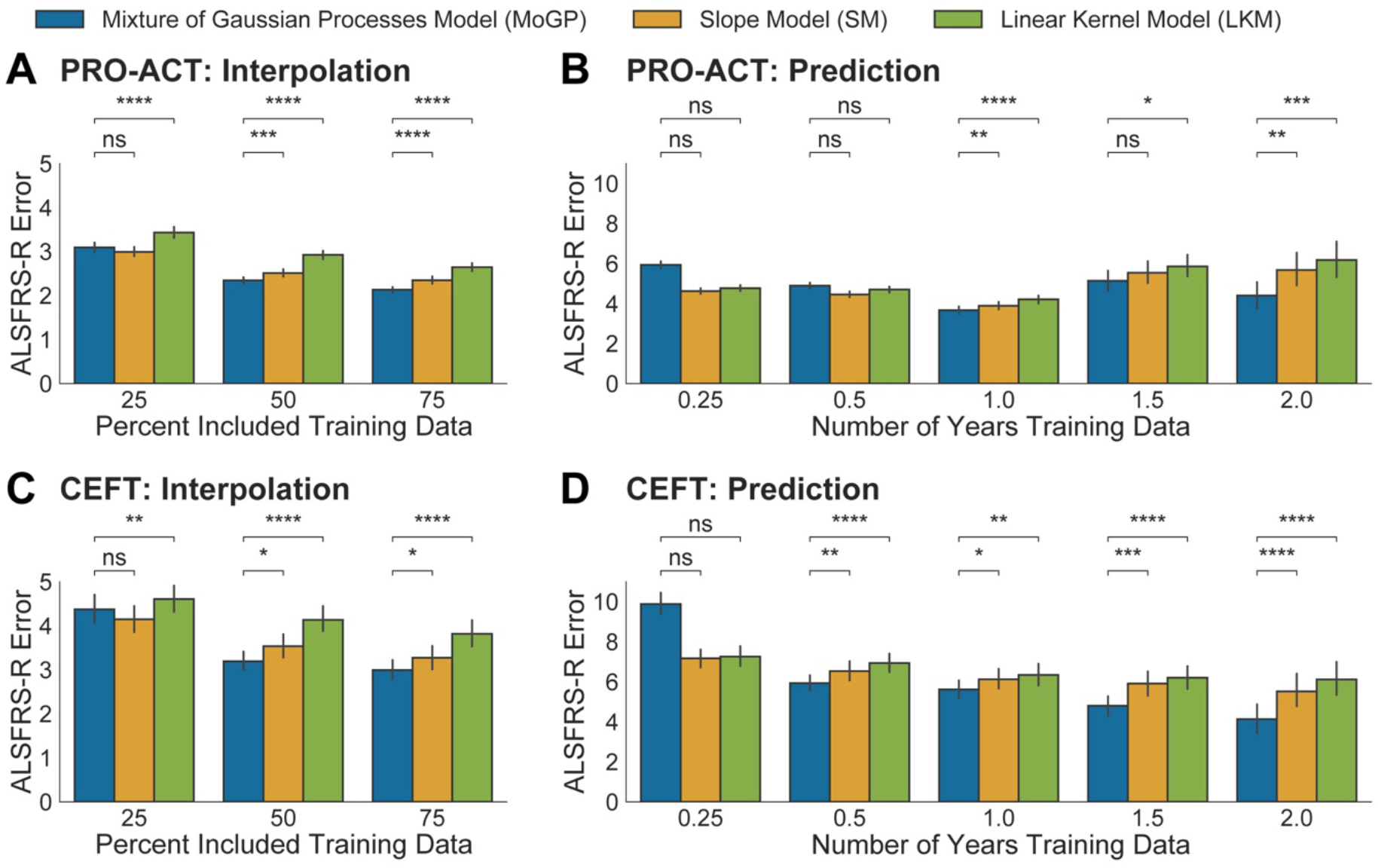
Evaluating robustness of cluster assignments with sparse datasets. (A,C) MoGP. LKM, and SM were trained on interpolated data and RMSE was calculated between withheld data and mean predicted trajectory (B D) Models were trained on right-censored training data *p-val ≤ 1e-1 **p-val ≤ 1e-2 ***p-val ≤ 1e-3 ****p-val ≤ 1e-4 (Wilcoxon signed-rank one-sided test). Error bars show 0.95 confidence interval

One of the most common uses for trajectory modeling is to predict future ALSFRS-R scores, and in clinical trials, these predictions are often made with the Slope Model (SM), the patient-specific linear model. For PRO-ACT, when only three or six months of data are provided, the SM and LMK are the most accurate (Fig. 3B). However, when one or more years of training data were provided, the MoGP model outperformed the LKM and SM (Fig. 3B, *p-val* <= 1e-2, except for 1.5 years where *p-val* =1.3e-1 for SM), and more accurately predicted future disease progression by more than 0.22, 0.41, and 1.28 ALSFRS-R points at 1, 1.5, and 2 year time points respectively. This trend was strengthened in CEFT, in which six months of training data was sufficient to see an improvement in progression forecasting (Figure 3D, *p-val* <=1e-1).

For the majority of comparisons, the MoGP identified fewer number of clusters per mixture model than the SM or LKM, indicating that the lower reconstruction error was not due to overfitting of the cluster assignments (Supplementary Fig 2).

### Trajectories are transferrable across all study populations

Because ALS is heterogeneous and characteristics of study populations can differ significantly, it is important that trajectory models capture signal that is consistent across populations. We evaluated the ability of MoGP, trained on a large database as a reference model, to predict patient trajectories from participants in other study populations with varying data collection frequencies and follow-up periods.

We found that the reference model demonstrated strong performance on external datasets, indicating that the trajectory clusters are not overfitted to the reference model data (Fig. 4A). Importantly, we found that for all test datasets (AALS, CEFT, EMORY), the Reference Model outperformed the Study-specific models (Fig. 4B, *p-val* < 0.05). AALS had the lowest error when the Reference model was used (2.16 ALSFRS-R points), followed by CEFT (2.25), and EMORY (2.32) (Fig. 4B). These errors were similar to the Baseline Error (1.88 ALSFRS-R points), which was the error when the Reference Model (trained on 60% of the participants in the PRO-ACT dataset) was tested on the remaining 40% participants. These errors were all much lower than that of the randomized PRO-ACT control, which had a mean error of 11.74 ALSFRS-R points. If the reference model was overfit, we would expect that when exposed to different populations, the study-specific models would have a significantly lower error than the reference model. Given that CEFT is a subset of PRO-ACT, it is interesting that CEFT study-specific model has a higher error than its reference model counterpart; these results suggest that the larger size of the PRO-ACT dataset may allow it to capture trajectories more accurately. The reference model’s ability to outperform all of the study-specific models is strong evidence that the trajectory patterns identified by MoGP are transferrable across ALS study populations.

**Figure 4.**
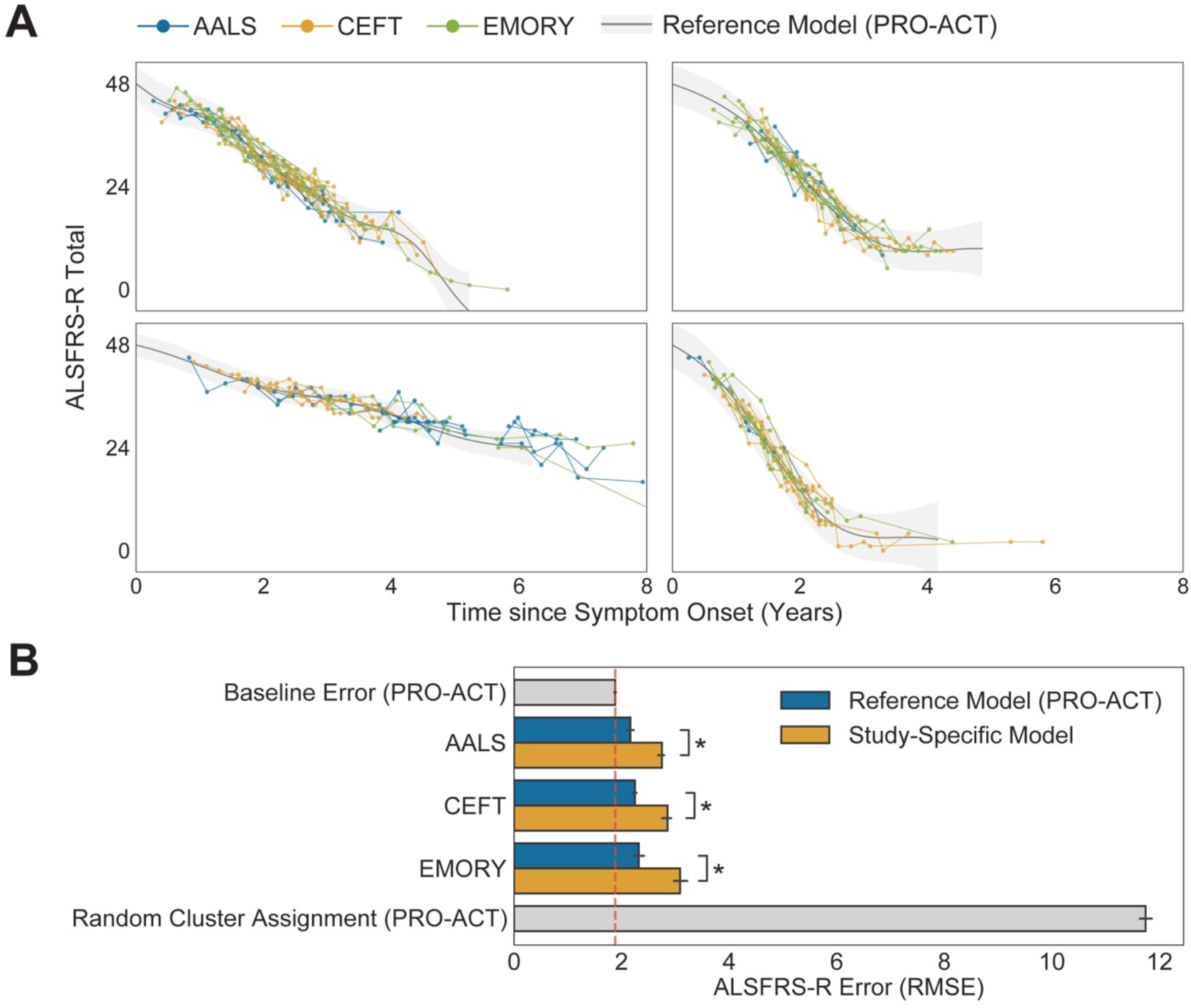
Assessing trajectory consistency across datasets. A) The reference model was trained on PRO-ACT and used to predict progression trajectories of participants in other datasets; the four largest reference model clusters are shown. B) Average test error between cluster mean function and participant ALSFRS-R scores, using the reference model and study-specific models, *p-val< 0.05 (Wilcoxon signed-rank two-sided test). Error bars show 0.95 confidence interval between 5 splits.

### Survival outcomes correspond with trajectory clusters

We next aimed to evaluate if the MoGP-identified clusters, which were trained only on ALSFRS-R data, were able to reflect the duration of patient survival from symptom onset to death. The results of the Kaplan-Meier analysis are presented in Figure 5. Some clusters (Fig. 5 C, E) reflected longer survival durations, with very few deaths recorded. Other clusters reflected shorter durations, corresponding to faster progression; cluster D has a median survival of 2.90 years from symptom onset (Fig. 5 D). Overall, we found that the MoGP trajectory patterns corresponded closely with survival probabilities, indicating clinical relevance of the identified ALS progression clusters.

**Figure 5.**
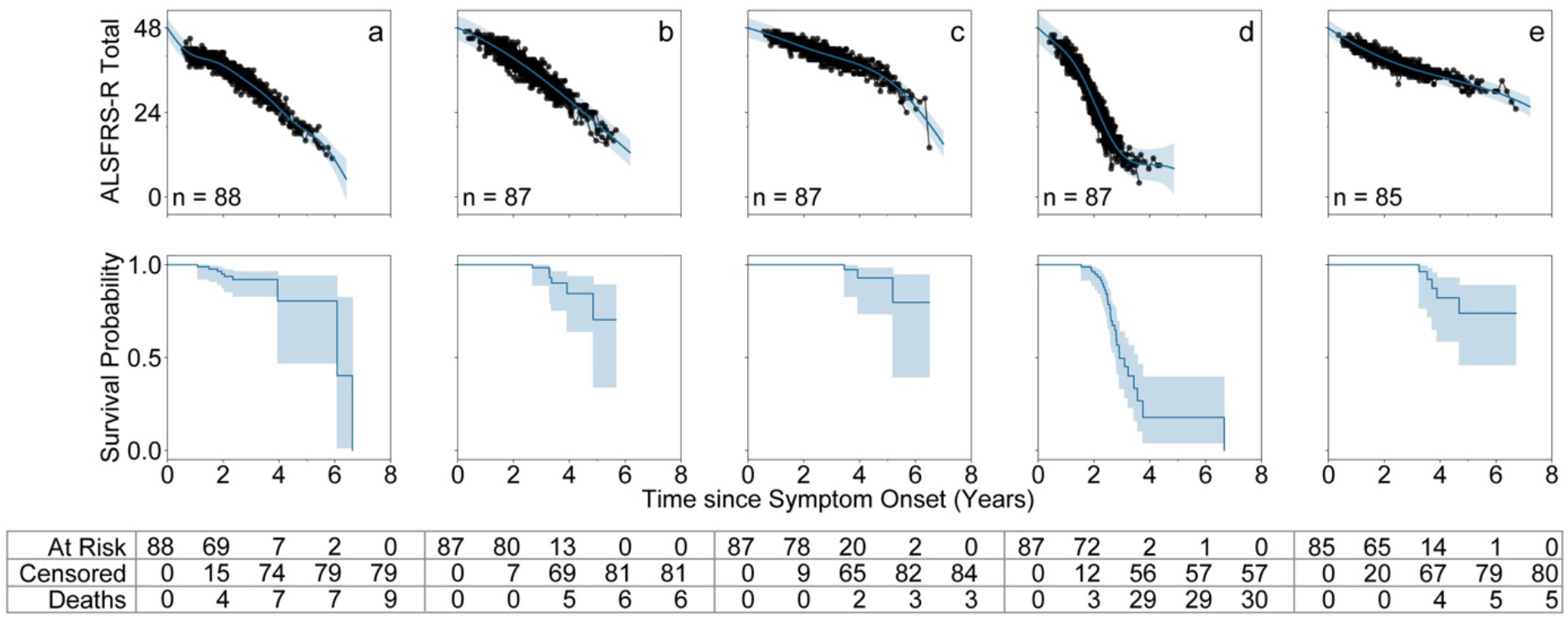
Survival outcomes for trajectory clusters. The five largest PRO-ACT clusters are shown with MoGP cluster (top) and cluster Kaplan-Meier survival curve (middle). The number of individuals at risk. censored, and with recorded deaths observed at each time point displayed as table (bottom).

### Common patterns of decline are found in forced vital capacity and ALSFRS-R subscores

In addition to ALSFRS-R scores, there are other important clinical metrics that can be used to monitor ALS disease progression. One is Forced Vital Capacity (FVC), which is a spirometer-based measure of lung function and has been used as an indicator of survival and disease progression.^33^ Furthermore, while ALSFRS-R Total is commonly used as an aggregate measure, its component subscores measuring fine motor, gross motor, bulbar, and respiratory function can also be analyzed to identify subscore-specific patterns. When we applied MoGP to FVC and ALSFRS-R subscores from PRO-ACT, we saw that the nonlinearity persisted in these domains as well. The nonlinear trajectories were particularly pronounced for FVC and bulbar function (Fig. 6). Overall, MoGP enables flexible trajectory identification from longitudinal metrics.

**Figure 6.**
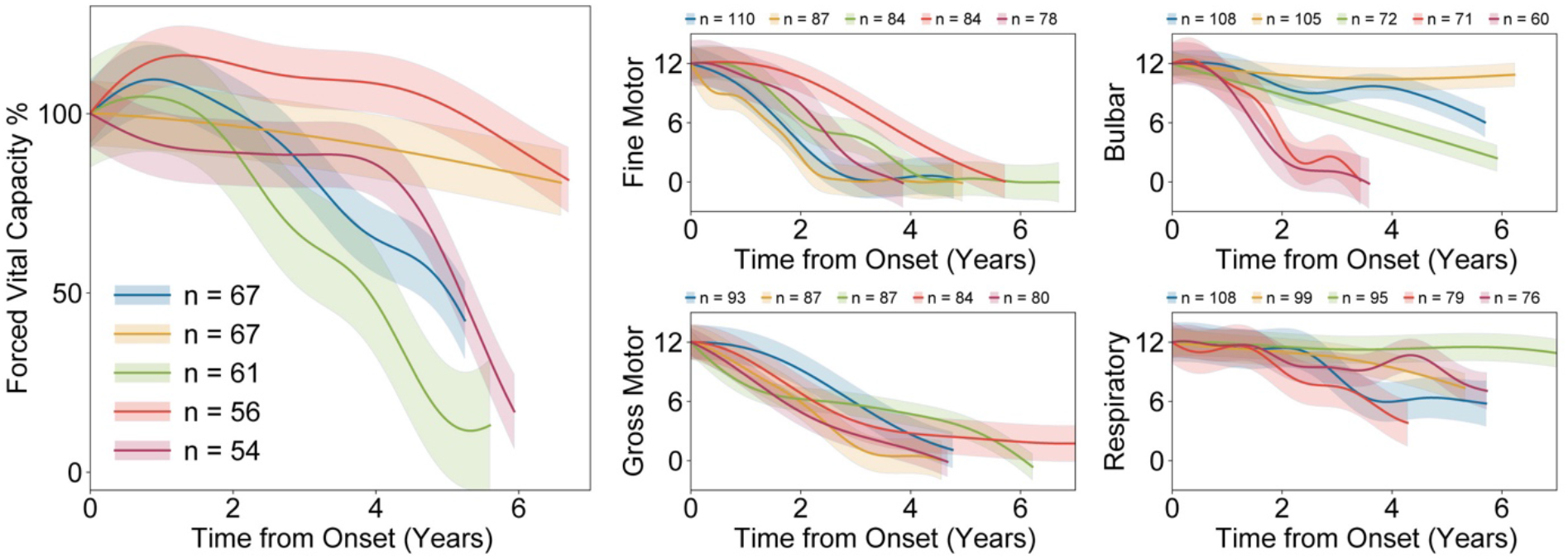
MoGP trajectory patterns for secondary endpoints of ALS disease progression: Forced Vital Capacity and ALSFRS-R subscores (fine motor, gross motor, bulbar, respiratory domains). Trajectory colors correspond to the relative number of participants in each cluster, scaled from largest to smallest of five largest clusters from PRO-ACT. Participants with minimal change in score (51 point) were excluded from the model

## Discussion

We characterize disease progression in ALS using a framework for identifying trajectory patterns from longitudinal data. While previous work on disease progression modeling has focused on patient-specific prediction models^16,34,35^, a critical advance of this work is the characterization of distinct and large trajectory clusters. Furthermore, we provide a principled approach to characterizing the shapes of disease progression patterns in ALS, which leverages Bayesian nonparametric methods to minimize the number of assumptions that are required for regression models.

The improved performance of a MoGP model over the slope and linear kernel models indicates that linear models are insufficient to capture the heterogeneity in ALS disease progression. While some patients do indeed have linear trajectories, a significant portion of patients have non-linear trajectories. Furthermore, the MoGP model does not prevent the discovery of clusters with primarily linear trajectories. Previous work has suggested that “functional cliff” patterns seen here may be a result of inconsistencies in the ALSFRS-R or issues related to the ordinal scale used in ALSFRS-R as opposed to a linearly-weighted interval scale.^36,37^ However, the consistency of MoGP-identified patterns across different study populations and different metrics (ALSFRS-R and vital capacity scores) instead point to nonlinear changes in patient function that are not solely dependent on clinical metric.

Our work also demonstrates how existing clinical databases in ALS can be leveraged to enable characterization of disease progression models in sparse datasets from different study populations. A MoGP model trained on the PRO-ACT database accurately predicted trajectories for clinical datasets from AALS, EMORY, and CEFT datasets. The transferability of MoGP-identified clusters across datasets indicates that the trajectory cluster patterns are robust to batch effects due to clinician or site differences.

These findings have implications for clinical trials designs, many of which use ALSFRS-R and vital capacity metrics as primary or secondary endpoints. In many trajectory clusters, functional cliffs or sigmoidal patterns in disease progression may obscure detection of therapeutic efficacy if linear models are used. While short clinical trials may not benefit significantly from using complex nonlinear models, for clinical trials that are one year in duration, or those with open label extension, MoGP can provide more accurate trajectory models. For PRO-ACT, we saw a reduction of error of 1.19 ALSFRS-R points with two years of training data. In context, clinical trial efficacy of edaravone was FDA approved with a 2.5 point difference in decline over 6 months^14^, and the efficacy of Sodium Phenylbutyrate–Taurursodiol was demonstrated by a difference in 0.42 points per months over six months, which approximates to a 2.52 point difference on the ALSFRS-R scale.^12^

MoGP can also be used to inform clinical trial participant stratification^5,38^ in cases where there are reliable pre-trial clinical data. If those data are available, the method can be used to stratify patients even for short clinical trials. Used prospectively, we would expect this approach to improve clinical trial statistical power.

Ultimately, by identifying clusters of patients that have similar disease progression trajectories, these models could be used to identify molecular correlates that may be associated with ALS progression subtypes. While this work focuses on ALSFRS-R and vital capacity, the field of ALS has identified a growing number of molecular biomarkers and clinical metrics in which progression is poorly understood.^39,40^ This paper points to the complexity of disease progression in ALS and the necessity of more accurately accounting for heterogeneous trajectory patterns in clinical trials models and research studies.

## Data Availability

We provide the python code for the MoGP framework as well as the pre-trained reference model described here for researchers to use to generate predictions of cluster membership and trajectory function from input patient data. All code used for data processing, modeling, and figure generation can be found at: https://github.com/fraenkel-lab/mogp
AALS is publicly available for download (data.answerals.org). PRO-ACT can be downloaded by request (https://nctu.partners.org/ProACT). CEFT can be downloaded from National Institute of Neurological Disorders and Stroke (NINDS) (https://www.ninds.nih.gov/Current-Research/Research-Funded-NINDS/Clinical-Research/Archived-Clinical-Research-Datasets) by request. EMORY is restricted access at this time.

https://data.answerals.org

https://nctu.partners.org/ProACT

https://www.ninds.nih.gov/Current-Research/Research-Funded-NINDS/Clinical-Research/Archived-Clinical-Research-Datasets

## Abbreviations

AALS: Answer ALS
ALS: Amyotrophic Lateral Sclerosis
ALSFRS-R: Revised ALS Functional Rating Scale
CEFT: Clinical Trial of Ceftriaxone in ALS
EMORY: Emory ALS Clinic database
FVC: Forced Vital Capacity
LKM: Linear Kernel Model
MoGP: Mixture of Gaussian Processes model
PRO-ACT: The Pooled Resource Open-Access ALS Clinical Trials
RMSE: Root mean squared error
SM: Slope Model

## Acknowledgements

ALS Finding a Cure and Packard Foundation supported the collection of the Answer ALS clinical data set used in the manuscript.

The Muscular Dystrophy Association (MDA) contributed funding to the Emory ALS Clinic database which was included in this research.

Data used in the preparation of this article were obtained from the Pooled Resource Open-Access ALS Clinical Trials (PRO-ACT) Database. As such, the following organizations and individuals within the PRO-ACT Consortium contributed to the design and implementation of the PRO-ACT Database and/or provided data, but did not participate in the analysis of the data or the writing of this report:

- Neurological Clinical Research Institute, MGH
- Northeast ALS Consortium
- Novartis
- Prize4Life Israel
- Regeneron Pharmaceuticals, Inc.
- Sanofi
- Teva Pharmaceutical Industries, Ltd.
- Knopp Biosciences

This research includes the National Institute of Neurologic Disease and Stroke’s Archived Clinical Research data (Clinical Trial of Ceftriaxone in ALS, Merit Cudkowicz, Massachusetts General Hospital) received from NINDS Archived Clinical Research Datasets webpage.

