## Supplemental Methods and Figures for "Identifying Patterns of ALS Progression from Sparse Longitudinal Data"

#### Study Populations

Data used in the preparation of this article were obtained from the Pooled Resource Open-Access ALS Clinical Trials (PRO-ACT) Database. In 2011, Prize4Life, in collaboration with the Northeast ALS Consortium, and with funding from the ALS Therapy Alliance, formed the Pooled Resource Open-Access ALS Clinical Trials (PRO-ACT) Consortium. The data available in the PRO-ACT Database has been volunteered by PRO-ACT Consortium members.

#### Modeling Approach

##### Gaussian Process Regression

Gaussian processes take the form:

$$f(x) \sim GP\ (m(x), k(x, x'))$$

where m(x) describes the model's mean function and k(x,x') describes the model's covariance function. To specify the covariance function, the Gaussian processes in this implementation of MoGP uses the SE kernel, with the form:

$$k(x, x') = \sigma^2 exp\ (-\frac{(x - x')^2}{2l^2})$$

where $\sigma^2$ is the signal variance and $l$ is the length-scale. The signal variance ($\sigma^2$) determines the average distance of the function from the mean. The length-scale ($l$) specifies the smoothness of the function, with increasing length-scales resulting in smoother functions. For the length-scale, a gamma prior with a mean of 4 and variance of 9 was used. The length-scale prior is approximately half of the maximum trajectory duration included in our model, selected to encourage minimal mean function crossings.

In contrast to the SE kernel, the LKM kernel is a linear kernel added to a bias kernel, with the form:

$$k(x, x') = \sigma_v^2 (x)(x') + \sigma_b^2$$

where $\sigma_v^2$ and $\sigma_b^2$ are the signal and bias variance, respectively. The bias allows for a non-zero intercept.

For both Gaussian Process kernels, a fixed signal variance of 1 was used to train the models. A gamma prior with mean 0.75 and variance of $0.25^2$ was used for the likelihood noise variance, selected to account for noise present in the data; this parameter was optimized through model training.

#### Dirichlet Process Clustering

This implementation of DP clustering uses a collapsed Gibbs sampler, in which we iteratively assign probabilities of each sample joining either existing clusters or generating a new cluster to calculate the likelihood of cluster membership. By repeating this process for each sample until convergence, we are able to identify the number of clusters the data captures as well as sample-specific cluster assignments.

The DP clustering model in MoGP takes the form:

$$G(f) = \sum_{k=1}^{\infty} \pi_k \delta_{f_k}(f)$$

where f indicates a cluster-specific GP regression function, k indicates cluster membership, and $\pi_k$ indicates cluster-specific probability, where:

$$\pi_k = \beta_k \prod_{j=1}^{k-1} (1 - \beta_j) \text{ and } \beta_k \sim Beta\,(\cdot\,|1, \propto)$$

$\propto$ indicates the scaling parameter and modifying this can influence the degree of cluster discretization and therefore the number of identified clusters.

#### Monotonic Inductive Bias

To encourage monotonically declining functions, we use two modifications to MoGP: 1) a negative linear mean function in our GP cluster, and 2) a thresholding function to determine cluster membership. In our thresholding function for each cluster, if the initial visit score for a given sample is above a threshold of the predicted mean score from the current cluster at the time of the initial visit, the probability of joining that cluster is set to 0. This prevents the probability that a participant with a starting ALSFRS-R score vastly divergent from a given cluster will be added to the cluster. For our negative linear mean function, we used a gamma prior with a mean of 0.66 and variance of 0.2. Together, these values were chosen to minimize major deviation from a monotonic trajectory, although these are weak priors that allow for a degree of non-monotonic behavior depending on the data.

### Supplemental Tables and Figures

| Dataset | Inclusion Criteria | Total No. Participants Included | No. Visits Mean (SD) | Months Followed Mean (SD) | ALSFRS-R Slope Mean (SD) |
|---|---|---|---|---|---|
| PRO-ACT | ≥ 4 visits | 2814 | 10 (4) | 12.18 (6.24) | -1.05 (0.87) |
| CEFT | ≥ 4 visits | 453 | 10 (5) | 19.79 (10.35) | -1.15 (0.71) |
| PRO-ACT | ≥ 10 visits | 1327 | 14 (3) | 15.55 (6.70) | -0.89 (0.65) |
| CEFT | ≥ 10 visits | 228 | 14 (4) | 26.91 (9.21) | -0.81 (0.43) |

**Supplement Table 1.** Study population summary statistics for participants included in prediction (≥ 4 visits) and interpolation (≥ 10 visits) experiments

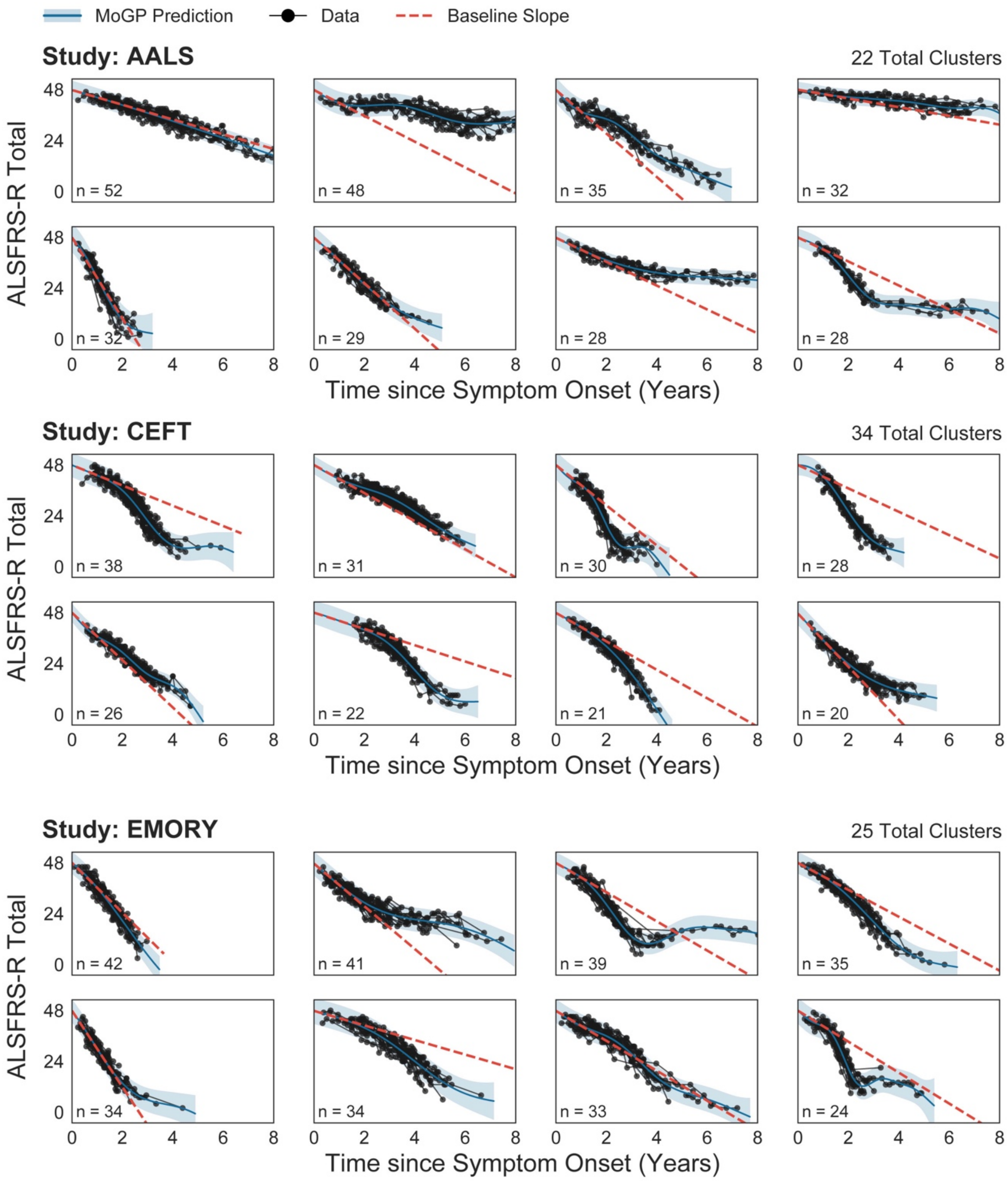


**Supplement Figure 1. Eight largest clusters for AALS, CEFT, and EMORY study populations visualized.** The baseline slope is calculated as the difference between 48 and the mean cluster score one year after symptom onset. N indicates the number of ALS patients in each cluster.

| | RMSE Difference (ALSFRS-R points) | | | | | |
|---|---|---|---|---|---|---|
| **Study Population** | > 0 | > 1 | > 2 | > 3 | > 4 | > 5 |
| **AALS** | 71.27% | 27.19% | 8.33% | 2.85% | 0.88% | 0.22% |
| **CEFT** | 75.42% | 39.29% | 16.18% | 6.51% | 3.78% | 1.26% |
| **EMORY** | 74.44% | 31.08% | 11.78% | 5.01% | 2.01% | 1.50% |
| **PRO-ACT** | 77.87% | 27.16% | 9.99% | 3.73% | 1.27% | 0.31% |

**Supplement Table 2: Percentage of patients that have a reduction in error when MoGP is used as compared to LKM.** ALSFRS-R point thresholds range from 0 ALSFRS-R points (indicates percent of all participants for whom a MoGP provides predictions with a lower error than LKM) to 5 ALSFRS-R points (percentage of patients that have a reduced error of more than 5 ALSFRS-R points using a MoGP).

| | RMSE Difference (ALSFRS-R points) | | | | | |
|---|---|---|---|---|---|---|
| **Study Population** | > 0 | > 1 | > 2 | > 3 | > 4 | > 5 |
| **AALS** | 37.28% | 11.40% | 3.29% | 1.32% | 0.44% | 0.00% |
| **CEFT** | 59.24% | 24.37% | 6.30% | 2.73% | 2.10% | 0.42% |
| **EMORY** | 45.61% | 11.03% | 4.76% | 2.51% | 1.50% | 0.75% |
| **PRO-ACT** | 61.27% | 19.43% | 5.71% | 1.95% | 0.58% | 0.17% |

**Supplement Table 3:** Percentage of patients that have a reduction in error when MoGP is used as compared to SM.

| Study Population | No. RBF clusters | No. LKM clusters | No. slope models |
|---|---|---|---|
| **AALS** | 22 | 25 | 456 |
| **CEFT** | 34 | 44 | 476 |
| **EMORY** | 25 | 30 | 399 |
| **PRO-ACT** | 92 | 127 | 2923 |

**Supplement Table 4: Number of clusters in each of the models, across study populations.** Because a slope model was fit to each patient, the number of slope models is equivalent to number of patients included in the training data.

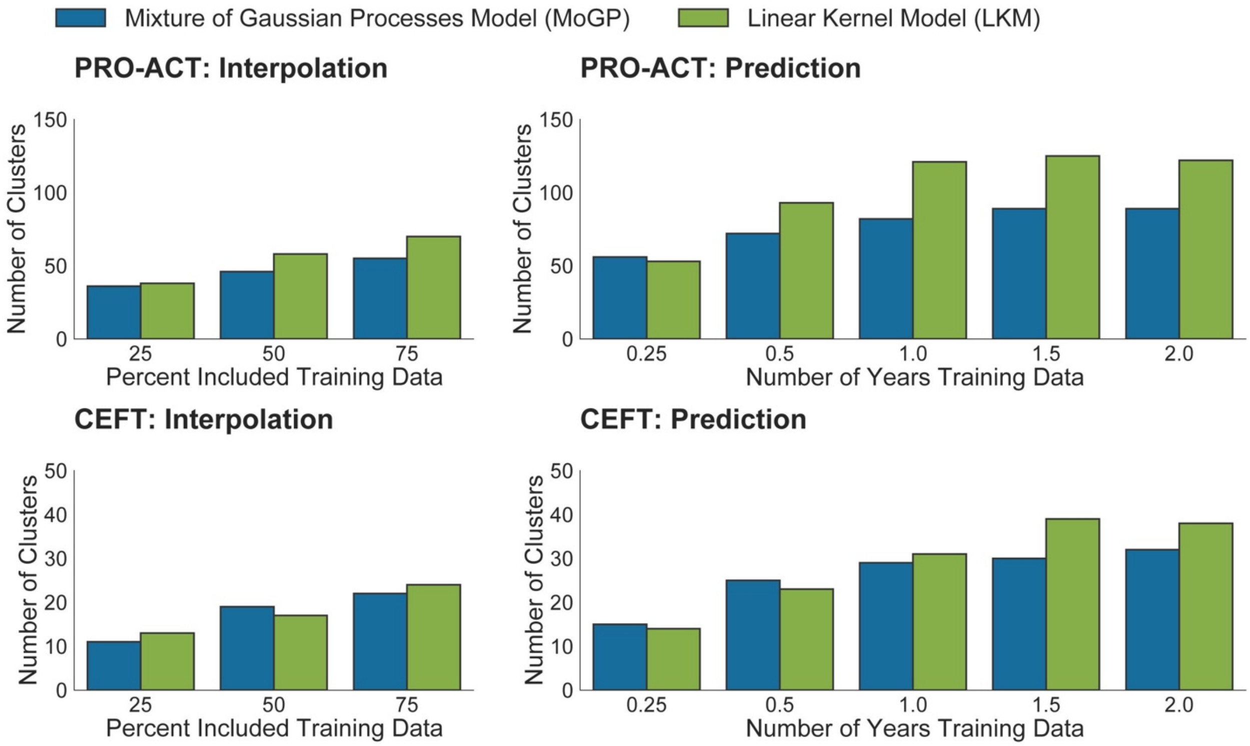


**Supplement Figure 2.** Number of clusters in MoGP and LKM models for interpolation and prediction tests, across PRO-ACT and CEFT datasets.
